# Image-localized biopsy mapping of brain tumor heterogeneity: A single-center study protocol

**DOI:** 10.1101/2022.11.14.22282304

**Authors:** Javier C. Urcuyo, Lee Curtin, Jazlynn M. Langworthy, Gustavo De Leon, Barrett Anderies, Kyle W. Singleton, Andrea Hawkins-Daarud, Pamela R. Jackson, Kamila M. Bond, Sara Ranjbar, Yvette Lassiter-Morris, Kamala R. Clark-Swanson, Lisa E. Paulson, Chris Sereduk, Maciej M. Mrugala, Alyx B. Porter, Leslie Baxter, Marcela Salomao, Kliment Donev, Miles Hudson, Jenna Meyer, Qazi Zeeshan, Mithun Sattur, Devi P. Patra, Breck A. Jones, Rudy J. Rahme, Matthew T. Neal, Naresh Patel, Pelagia Kouloumberis, Ali H. Turkmani, Mark Lyons, Chandan Krishna, Richard S. Zimmerman, Bernard R. Bendok, Nhan L. Tran, Leland S. Hu, Kristin R. Swanson

## Abstract

Brain cancers pose a novel set of difficulties due to the limited accessibility of human brain tumor tissue. For this reason, clinical decision-making relies heavily on MR imaging interpretation, yet the mapping between MRI features and underlying biology remains ambiguous. Standard (clinical) tissue sampling fails to capture the full heterogeneity of the disease. Biopsies are required to obtain a pathological diagnosis and are predominantly taken from the tumor core, which often has different traits to the surrounding invasive tumor that typically leads to recurrent disease. One approach to solving this issue is to characterize the spatial heterogeneity of molecular, genetic, and cellular features of glioma through the intraoperative collection of multiple image-localized biopsy samples paired with multi-parametric MRIs. We have adopted this approach and are currently actively enrolling patients for our ‘Image-Based Mapping of Brain Tumors’ study. Patients are eligible for this research study (IRB #16-002424) if they are 18 years or older and undergoing surgical intervention for a brain lesion. Once identified, candidate patients receive dynamic susceptibility contrast (DSC) perfusion MRI and diffusion tensor imaging (DTI), in addition to standard sequences (T1, T1Gd, T2, T2-FLAIR) at their presurgical scan. During surgery, sample anatomical locations are tracked using neuronavigation. The collected specimens from this research study are used to capture the intra-tumoral heterogeneity across brain tumors including quantification of genetic aberrations through whole-exome and RNA sequencing as well as other tissue analysis techniques. To date, these data (made available through a public portal) have been used to generate, test and validate predictive regional maps of the spatial distribution of tumor cell density and/or treatment-related key genetic marker status to identify biopsy and/or treatment targets based on insight from the entire tumor makeup. This type of methodology, when delivered within clinically feasible time frames, has the potential to further inform medical decision-making by improving surgical intervention, radiation, and targeted drug therapy for patients with glioma.

## Introduction

Brain tumors are uniquely challenging due to the limited access intrinsic to their location delaying progress in understanding and developing effective treatments. For example, the most aggressive primary brain cancer, glioblastoma (GBM), is particularly well known for its short median survival from diagnosis, and once systemic cancers reach the stage of brain metastases, patients typically experience very poor outcomes [1–3]. Surgery remains one of the most notable treatments for all brain tumors. In some cases, the benefits go past debulking, as surgery also provides an opportunity to collect tissue and elucidate genetic, transcriptomic and environmental factors that may act as treatment targets or prognostic markers. However, capturing the full heterogeneity within brain tumors remains impossible given the eloquent brain tissue in which these tumors live. The protocol introduced is intended to lead to a large dataset of image localized biopsies that will be able to answer some of these key questions surrounding the heterogeneity of brain tumors.

The landscape of each brain tumor is ever changing, and there are limited opportunities for tissue collection, so it is vital that these limited surgical sampling opportunities are capitalized on. Diffusely invasive gliomas are particularly challenging in this regard as there are always tumor cells left behind in the brain following any surgical intervention. GBM, the most aggressive form of glioma, is the most common primary brain malignancy among adults [4]. Despite aggressive medical intervention consisting of maximal surgical resection followed by concurrent chemoradiation and adjuvant temozolomide, this disease remains uniformly fatal, with a median survival rate of 14-16 months in patients with newly-diagnosed GBM [5,6] and 5-7 months in patients with recurrent GBM [7]. This poor prognosis is often a result of a major hallmark of GBM: profound intratumoral heterogeneity that contributes to treatment resistance and tumor recurrence. The presence of metastatic brain lesions also corresponds to a poor prognosis. These lesions have common alterations compared to their primary counterparts [8]. However, there is also some evidence pointing towards diversity within and between metastatic brain lesions [9,10]. We collect metastatic tumor tissue to understand this heterogeneity and also determine how the different metastatic tissue features may present on imaging.

Given the nexus of known intratumoral heterogeneity and the limitations of access to brain tumor tissue, there is an urgent need to leverage imaging to better inform our understanding of the biology at play across patients and within each patient’s tumor. For example, the molecular composition of tumors is important, as different tumor cell subpopulations within and between patients can have different treatment sensitivities and implications for survival. For instance, IDH1/IDH2 and MGMT are prognostic markers of survival for gliomas [5,6]. Notably, mutation in IDH is thought to be uniform throughout a patient tumor [11,12], while MGMT status can vary with treatment status and location [13–15]. Other genetic alterations can arise, with multiple phenotypes present within the same tumor. For instance, the epidermal growth factor receptor (EGFR) gene can differ significantly in different regions of the tumor [16–19]. Similarly, EGFR-targeted therapies have mixed responses in patients [20,21]. Bulk transcriptional studies of GBM, derived from The Cancer Genome Atlas data, have revealed tumor subtypes that are associated with specific genetic alterations and patient survival [22,23]. These subtypes have been recapitulated on the level of individual cells, and profound genomic and phenotypic variability has been identified within individual patients [24–27]. These heterogeneities in key markers suggest that, although sufficient for diagnosis, clinical samples likely do not represent the full genetic and transcriptomic complexity of each tumor. It is important to have an accurate representation of the tumor throughout the clinical course of care, but opportunities for tissue collection are limited. This lack of tissue access has led to a heavy reliance on imaging to assess important clinical benchmarks such as surgical outcome, tumor size, and treatment response.

Due to its ubiquitous use in the clinic, there has been a long history of using MRI to predict the extent and biological heterogeneity of brain tumors [28–42]. Although they have shown promise, validating these predictions has been extremely difficult without access to local tissue samples. Image-localized biopsies, such as those collected here, have the potential to be used in developing and validating such spatial mathematical models as well as emboldening newer approaches such as radiomics, where machine-learning models are trained to connect high throughput quantitative image features to the composition of brain tumor tissue [17,19,43–48]. Given limited access to brain tumor tissue, typical radiomics studies aim to connect image features from tumors to global characteristics of the tumors such as malignancy, IDH1 status and grade [49] whereas image-localized biopsies provide an opportunity to embrace intratumoral heterogeneity. This same quantitative approach (image analysis meets tissue analysis meets machine learning) can and has been used to link local imaging patterns at biopsy sites with the composition of GBM tissue of said biopsies [17,19,43,47,48]. Given that many clinical decisions in the treatment and monitoring of GBMs are based on MRI, connecting GBM tissue composition to MRI offers the potential to capture the evolving tumor landscape and cellular populations noninvasively, leading to more informed clinical decisions to benefit the patient. As we collect more image-localized biopsies, this dataset becomes more representative of the vast range of possible tumor compositions and imaging appearances, giving machine learning models the opportunity to become more robust. Such robust models have the potential to predict important tissue features of GBM through imaging alone on a patient-specific spatial basis, which would arm clinicians with the knowledge to provide more nuanced treatment and better stratify patients for clinical trials.

This clinical protocol’s sample collection began in October 2017, and is ongoing. Collection of samples will continue in efforts to grow statistical power as long as we are able to conduct this research. This protocol has no impact on consented patients’ clinical care, nor alters anything within the clinical milieu.

### Imaging-defined tumor regions drive clinical decisions

Currently, maximal safe surgical resection is the clinical gold standard. However, this is often determined with postoperative magnetic resonance imaging (MRI), particularly the T1-weighted MRI with gadolinium contrast (T1+C) [50,51]. Tissue sampling creates significant challenges for studying the clonal diversity of tumors such as GBM. T1+C MRI represents the clinical standard for neuronavigation and routinely guides surgical biopsies and resection from the MRI enhancing core. In fact, a clinically defined “gross total resection”, for high grade glioma, is defined as removal of the complete T1+C abnormality. Unfortunately, biopsies from this contrast-enhancing (CE) tumor region alone fails to address the diverse molecularly-distinct subpopulations that extend into the surrounding non-enhancing (NE) parenchyma, which is visible on T2-weighted/Fluid-Attenuated Inversion Recovery (T2W/FLAIR) MRI [17]. These generally unresected NE tumor regions contribute to tumor recurrence and can have different cellular compositions and genetic signatures to that of enhancing regions [27,52,53]. Furthermore, T1+C MRI fails to localize cancer in the surrounding NE tumor region during radiation treatment (RT) planning, as non-tumoral edema typically appears visually indistinguishable from NE tumor. Most radiation oncologists must apply submaximal doses across the entire T2W/FLAIR volume, which delivers unnecessary radiation to the normal brain and risks undertreating NE tumor.

### Advanced multiparametric imaging provides deeper insights into tumor biology

Imaging techniques, such as advanced MRI, can quantitatively characterize tumor-induced physiological processes in the NE region of GBM. Unlike surgical sampling, MRI captures the entire brain organ and thus could provide insight into the tumor extent even in unresected NE regions and their associated biophysical features that may be detectable on MRI (Fig 1). On T1+C MRI, enhancement indicates regions of disrupted blood brain barrier (BBB), while signal demarcates regions of high water content and tumoral edema in T2W/FLAIR. Other advanced MRI features may reflect tumor cell density on diffusion-weighted imaging (DWI) [54], white matter infiltration on diffusion tensor imaging (DTI) [55,56], and microvessel morphology on Dynamic Susceptibility-weighted contrast-enhanced perfusion MRI (DSC-pMRI) [57]. In addition, signal intensity values on structural MRIs are spatial representations of soft tissue anatomy. The textural patterns of neighboring voxel intensities provide further insight towards the potential tissue microstructure and phenotypic heterogeneity within the local microenvironment [58,59]. These complementary MRI features offer potential biomarkers of underlying genomic and transcriptomic status, and have been previously correlated with molecular profiles of GBM [17,60–67]. Further, quantification of the interactions amongst molecularly-distinct subpopulations, cellular subpopulation compositions and/or their diversity in the NE tumor region (often left behind following surgical interventions) can help improve future treatment strategies (such as adaptive therapy), under the realm of individualized oncology [68–70].

**Fig 1:**
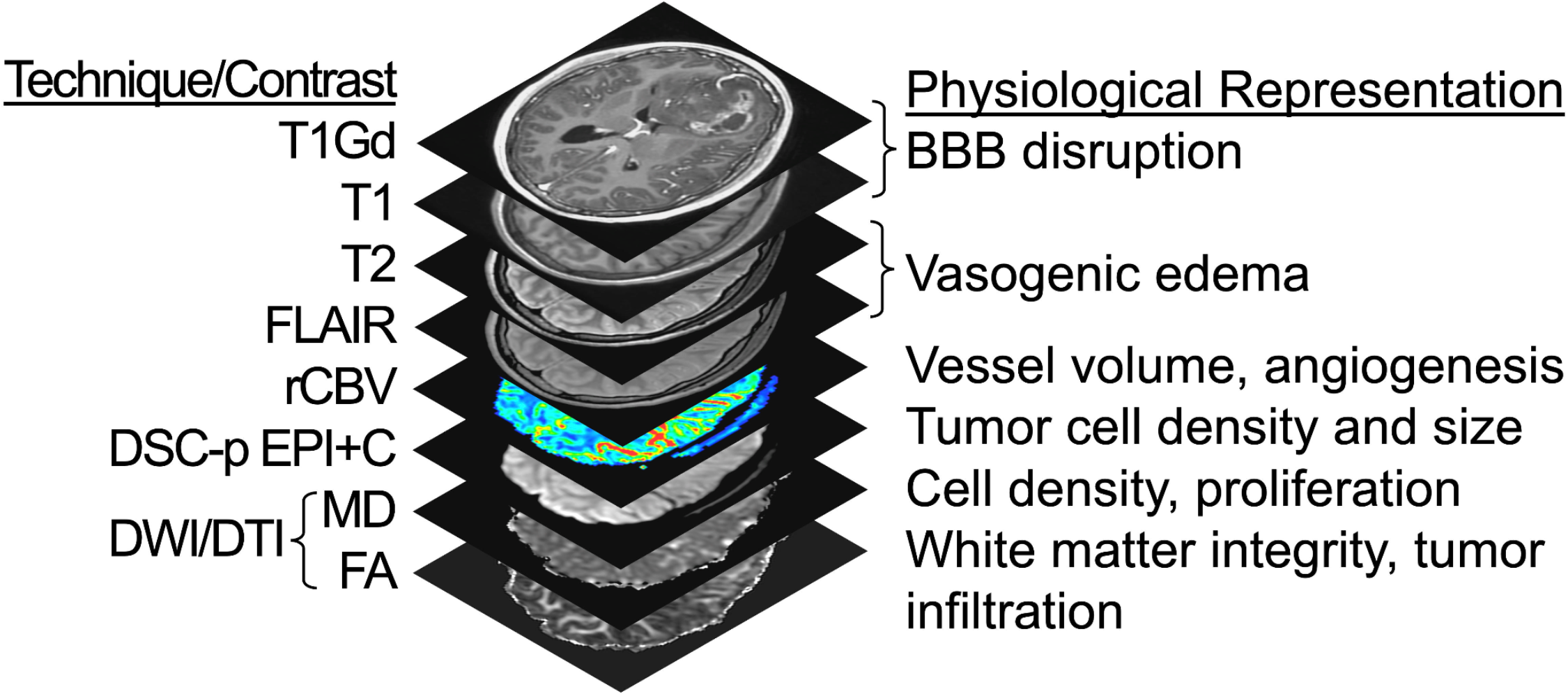
Multiparametric MRI techniques and contrasts. Listed are the 8 different MRI sequences used in this study, along with their corresponding physiological representations. T1Gd = T1W signal increase on post-contrast imaging; FLAIR = Fluid-Attenuated Inversion Recovery; rCBV = Relative cerebral blood volume; DSC-pMRI = Dynamic susceptibility contrast perfusion MRI; EPI+C = T2*W signal loss (i.e., negative enhancement at ∼5min post injection of 0.1 mmol/kg preload contrast injection); DWI = Diffusion weighted imaging; DTI = Diffusion tensor imaging; MD = Mean diffusivity (equivalent to apparent diffusion coefficient); FA = Fractional anisotropy.

## Materials and methods

The ‘Image-Based Mapping of Brain Tumors’ clinical protocol (IRB# 16-002424) is a minimal-risk study that aims to further characterize intratumoral heterogeneity of brain tumors. Here, we outline our study collecting multiple image-guided biopsy samples as part of standard surgical approaches. We aim to collect samples dispersed in and around the abnormality seen on T1Gd MRI to allow for cross-annotation for relevant MRI features. As these image-localized biopsies are collected during routine surgical interventions, this study does not alter the course of treatment for consented patients, but we hope that findings using this data will ultimately help patients in the future. We present an example set of biopsy locations for a single surgery in Fig 2. The data gathered through this study will be useful for countless research directions. The study’s primary objectives are to identify relationships between imaging and a number of important tumor features, such as tumor cell density, genetic status, transcriptomic status, and molecular status. Secondary objectives include investigating associations amongst imaging, radiation dosimetry, tumor recurrence/treatment effect, and clinical outcome. We foresee the use of statistical methods such as repeated measures correlations [71] and mixed effects models [72] to test for gene correlations and differences between imaging regions, Cox mixed effects models [73] to determine impact on clinical outcome, and supervised machine learning models such as regressors or classifiers to connect tumor composition (cellular, genetic, transcriptomics) to the more readily available imaging data.

**Fig 2:**
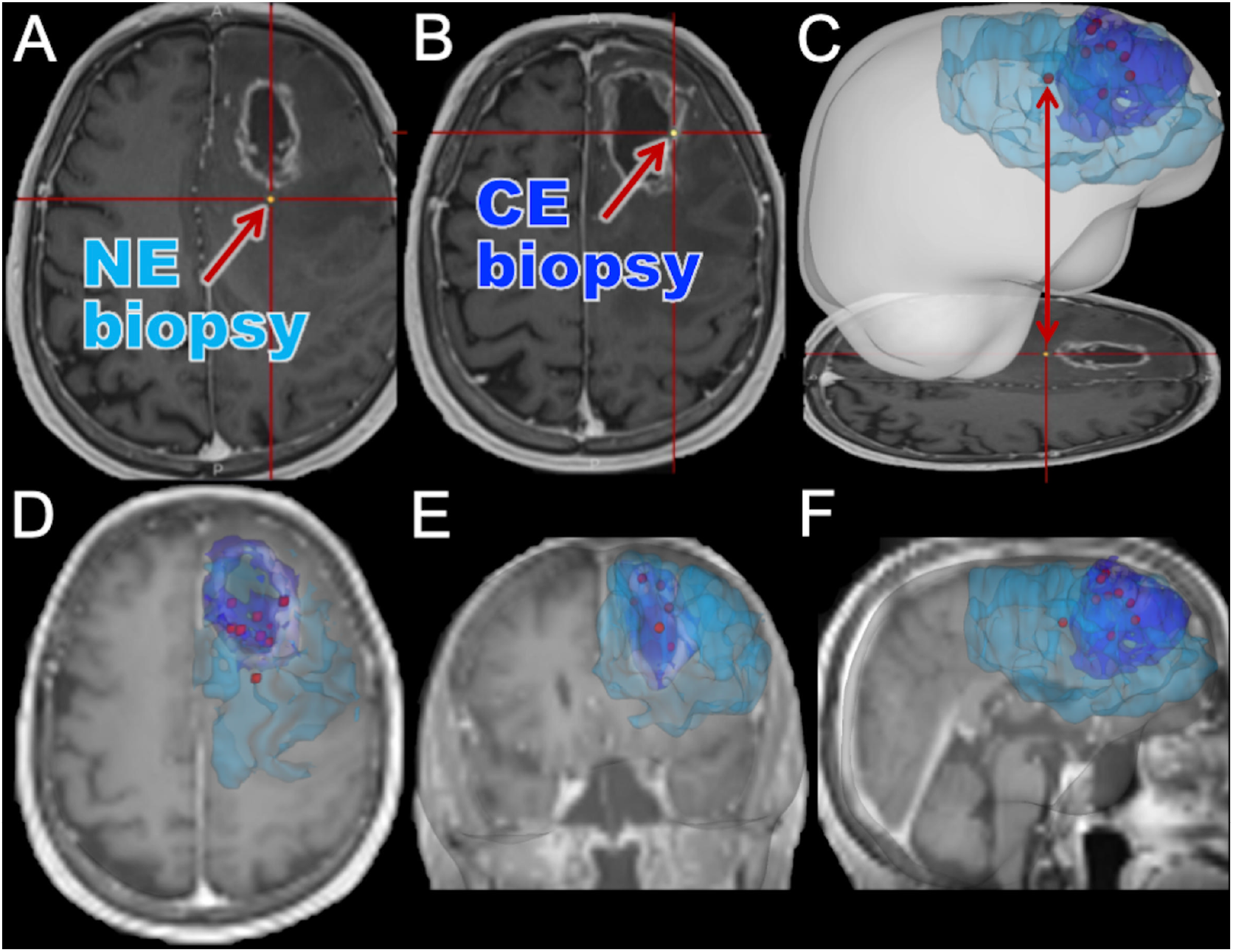
Illustrative example of biopsy locations annotated relative to imaging abnormalities. Here we show an example case where 9 image-localized biopsies were acquired from a patient. (A) An example screenshot from the intraoperative neuronavigation system of a non-enhancing (NE) biopsy sample. (B) An example screenshot from the intraoperative neuronavigation system of a contrast-enhancing (CE) biopsy sample. (C) A screenshot from intraoperative navigation is shown, highlighting the translation to a full three-dimensional rendering of segmented imaging abnormalities and biopsy locations. The segmented CE (dark blue) and NE (light blue) abnormalities are shown with biopsy locations (red). (D-F) MRI planes overlaid with segmentations and biopsy locations, highlighting the representative spread of biopsy locations that we can achieve. Although only the CE and NE regions are outlined here, we collect other multiparametric MRIs as well for the patients as shown in Fig 1.

### Inclusion and exclusion criteria

Inclusion criteria for patient participation in the study are as follows:

- Age between 18 and 99 (inclusive)
- Undergoing diagnostic biopsy and/or surgical resection for a brain lesion

Exclusion criteria for patient participation in the study are as follows:

- Insufficient renal function: eGFR < 60 mg/min/1.72m^2^ [74]
- Allergy to Gadolinium (Gd)
- Pregnant or nursing
- History of hemolytic anemia or asthma
- Inability to obtain informed written consent

### Workflow

#### Before surgery

Patients are identified through the institutional electronic medical records system (i.e., Epic Systems). Clinical schedules for MR scanners and operating rooms are checked daily for eligible participants. A study-specific multi-parametric imaging protocol is acquired prior to surgery for eligible patients. The imaging protocol includes T1-weighted (T1W), T1-weighted with gadolinium contrast (T1Gd), T2-weighted (T2W), T2W-fluid-attenuated inversion recovery (T2-FLAIR), diffusion tensor imaging (DTI), and dynamic susceptibility contrast (DSC) perfusion MRI (Fig 1). These imaging sequences are acquired prior to the collection of image-guided biopsy samples and, in some instances, following the patient’s diagnosis or treatment. In addition to MRI, other imaging modalities may be collected and reviewed, these include computed tomography (CT) and positron emission tomography (PET) imaging data collected as part of clinical practice or in conjunction with other imaging-based protocols.

Blood is collected in up to two 10mL ethylenediamine tetraacetic acid (EDTA) tubes and the buffy coat is frozen. This blood collection is used for germline DNA comparisons to the respective tumors and occurs during a standard of care visit. If the patient has multiple standard-of-care lab visits, we may collect at any of those as long as the subject has not withdrawn informed consent.

Urine is collected and stored at 4°C to assess biomarkers (free circulating DNA/RNA) associated with therapy response. This may also be collected at any standard of care lab visits, as long as the subject has not withdrawn informed consent. Table 1 shows the timeline for the collection of all sections of this protocol.

**Table 1:** General schedule of events for this research study from patient identification to routine follow-up. Patient information beyond routine follow-up can also be collected. No additional modifications are made to the standard clinical care trajectory, unless the patient has another surgery.

| Image-Localized Glioma Biopsy Protocol | Screening and Baseline | Day of Surgery | Post-Surgery follow-up | Routine Follow-up Visit |
| --- | --- | --- | --- | --- |
| Schedule of Events | Up to 30 days before surgery | Day 0 | Days 1-5 | Days 30-45 |
| Review Inclusion/Exclusion | X |  |  |  |
| Obtained Informed Consent | X |  |  |  |
| Imaging Protocol |  |  |  |  |
| T1 with and without contrast | X |  | X | X |
| T2/FLAIR | X |  | X | X |
| DTI | X |  | X<sup>1</sup> | X<sup>1</sup> |
| Perfusion | X |  | X<sup>1</sup> | X<sup>1</sup> |
| Tissue Collection |  | X |  |  |
| Blood Collection | X |  | X | X |
| Urine Collection | X |  | X | X |
| Clinical Data Collection | X | X | X | X |
| Screenshot Collection |  | X | X<sup>2</sup> |  |

X<sup>1</sup> - At the discretion of the principal investigators
X<sup>2</sup> - If screenshots are not collected on the day of surgery

#### During surgery

During routine surgery, biopsies are obtained and the MRI location is recorded using an intraoperative neuronavigation system (Medtronic StealthStation 8, Minneapolis, MN) with additional screenshots taken (such as that in Fig 3) to further validate the coordinate location. At the beginning of this study, biopsies were frozen by the Surgical Pathology department. However, to reduce time to freezing of the tissue, flash-freezing was implemented in the operating room (OR) with Surgical Pathology as a backup, if necessary. This has significantly improved our success in achieving freezing tissue within 5 minutes of surgical extraction (S1 Fig). We collect multiple spatially-annotated stereotactic biopsies from the across the diversity of MRI-definable tumor regions in patients with glioma and metastatic brain tumors with a goal of 8 biopsies per surgery with a target of ≥250 mg/biopsy (see Fig 4). Tumor grade is assessed by a pathologist using corresponding clinical biopsy samples in line with standard conventions [80,81].

**Fig 3:**
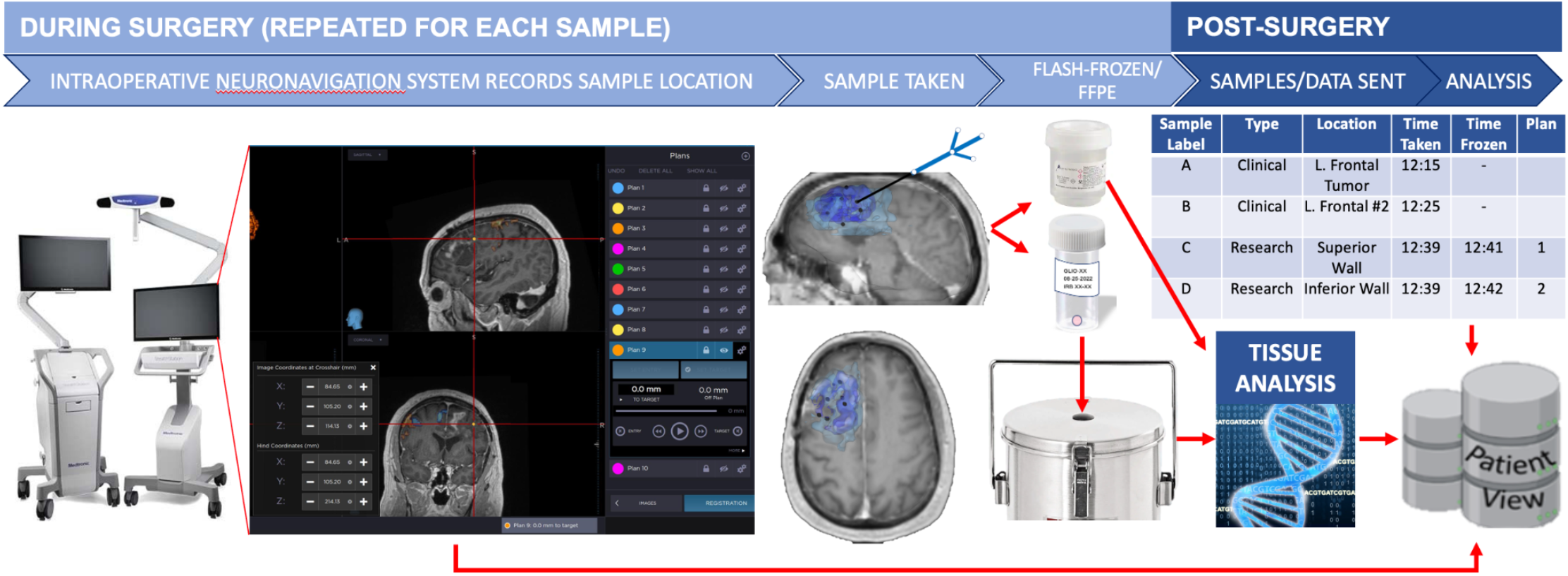
A schematic of our per-patient biopsy collection workflow. Additionally, patients may have samples collected at subsequent surgeries [75–79].

**Fig 4:**
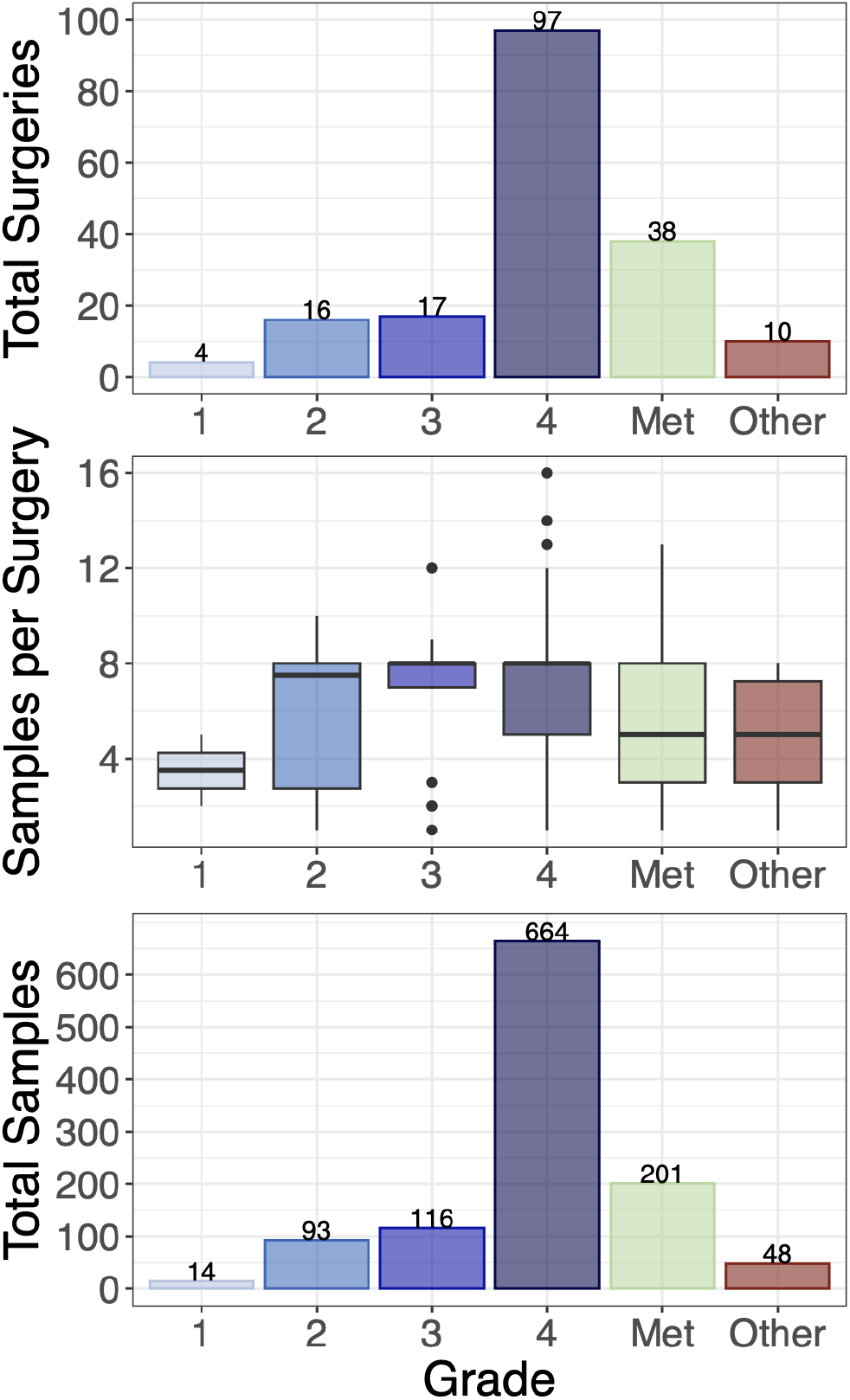
Overview of Image-Localized Biopsy Histology Distribution. **(Top)** Total number of surgeries with image-localized biopsies collected, broken down by grade (total of 183 surgeries). **(Middle)** Number of samples per surgery, broken down by grade. **(Bottom)** Total samples collected per grade (total of 1136 image-localized samples). Met = Metastatic brain tumor.

Originally, biopsies were only flash-frozen in order to conduct whole-exome and RNA sequencing on the samples. Since December of 2022, we also began submerging tissue in formalin. At first, this was done by the surgeon collecting extra samples, 2 at each coordinate location (16 formalin fixed samples were collected with this method). However, this overstretched the clinical workflow. In consultation with clinical staff, it was decided that one sample will be collected from each coordinate location, and if the sample is large enough the research team will split them in two; one is then flash frozen and the other formalin fixed. This way, we have more information about the biology in each of these areas, without any increased risk to the patient or workload on the clinical staff. We have so far collected a total of 29 formalin-fixed samples.

Screenshots taken for each research sample are extracted from the neuronavigation system and imported into our IRB-compliant database PatientView (Fig 3). Samples are time-stamped and designated an alphabetical letter to facilitate matching each sample to its corresponding screenshot and clinical notes in the electronic medical record. Samples collected are also cross-referenced with the Surgical Pathology department to confirm the number of biopsies collected, label accuracy, and overall quality assurance.

#### After surgery

Imaging data is collected, coded, processed, and matched with the genetic and molecular data obtained from each biopsy. Post-processing analyses include registration, normalization, inhomogeneity correction, and feature extraction as described elsewhere [17,43,48]. Planned studies include the development of statistical models between imaging, various tissue characterizations (e.g. copy number variants, transcriptomic signatures, immunohistochemistry etc.), and clinical outcomes.

Each sample is delivered to the Surgical Pathology department after being flash-frozen in liquid nitrogen. The samples are stored in a −80ºC freezer until subsequent processing. Flash-frozen tissue is retrieved and embedded frozen in optimal cutting temperature (OCT) compound. Tissue is sectioned (e.g. target 10μm with the goal of up to 20 slides) in −20ºC cryostat (e.g., Microm-HM-550) utilizing a microtome blade. Originally, In the event of excess archived formalin-fixed paraffin-embedded (FFPE) tissue collected per the standard clinical protocol, retrospective tissue may be obtained (under the discretion of a neuropathologist) to undergo further tissue analysis (we aim to collect up to 100μm in FFPE scrolls). Now, if size allows, samples are split by the research team and formalin-fixed in the OR. All specimens are also stained with hematoxylin and eosin (H&E) and reviewed by a neuropathologist to quantify tumor content. Tissue specimens may be submitted for subsequent genetic, molecular, or epigenetic analysis, including, but not exclusive to, next-generation sequencing, array-based comparative genomic hybridization (aCGH), exome sequencing, methylation analysis, and RNA sequencing.

### Data abstraction, management, and availability

Details of the patient’s clinical course, treatments, MRI images, pathologies, and treatment response are abstracted from the medical records system by IRB-approved staff and an anonymized data repository is distributed to the rest of the team for data analysis. Data is made accessible to all IRB-approved staff, but only the principal investigators, study coordinators, and research assistants have full rights to update data in our main data repository as they are responsible for ensuring the data quality and accuracy.

Biopsy-related data, including time of collection, specimen ID, neuronavigation information, and other notes are collected by two to three researchers during surgery in physical journals. These journals contain coded patient IDs with no patient health information. This data is transferred to a password-protected document with all relevant data for each patient, which is stored on a secure server that can only be accessed by IRB-approved researchers. Biopsy location data and image information is taken directly from the neuronavigation system using plans and time-stamped screenshots as previously described. The image data and screenshots are kept in deidentified patient folders on the secure server. Patient identifiers, demographic information, and sample data are collated in the main data repository.

Research data that documents, supports, and validates research findings will be made available after the main findings from the final research data set have been accepted for publication. We will provide these in the form of csv files. In addition, if requested, tissue and imaging data may be made available for sharing to qualified parties by the technology transfer office as soon as is reasonably possible, so long as such a request does not compromise intellectual property interests, interfere with publication, invade subject privacy, or betray confidentiality. Data that are shared will include standards and notations needed to interpret the data, following commonly accepted practices. All data from tissue processed from this cohort will be made available through the MOSAIC consortium we have formed around this important novel intratumoral heterogeneity initiative: www.BrainTumorMOSAIC.org. Additional data requests may be initiated by contacting the Swanson lab through our website: www.MathematicalNeuroOncology.org.

### Ethical considerations and declarations

This study was approved by the Mayo Clinic IRB on 1/9/2017 with approval number 16-002424. All participants presented in this work have undergone written informed consent. The principal investigators are responsible for ensuring the IRB-approved study protocol is followed and for reporting any adverse events. The study protocol is reviewed for renewal annually.

### Safety considerations

Our study is not therapeutically interventional. This is a minimal-risk study as procedures are in line with standard clinical activities with only the possible addition of advanced imaging. The safety of the patient is our top priority and is monitored by the neurosurgeon as surgery is performed.

### Patient demographics and sample counts

As of March 31st 2023, this study has enrolled 203 patients with 183 surgeries that resulted in the successful collection of biopsy samples (182 of which collected image-localized biopsies). A total of 1171 biopsies have been collected of which 1136 are image localized, with a mean of 6.40 samples per surgery. The mean age of these patients was 57.5 years old (ranging from 18-91). The self-reported sex breakdown is 113 males and 90 females with mean ages of 58.2 years (20-84) and 56.6 years (18-91), respectively. The self-reported racial and ethnic distribution of the cohort is largely white and non-Hispanic/Latino individuals (Fig 5, analogous sex-specific figures in Supplement 1). The samples collected in this study were largely acquired from patients with glioma and brain metastasis, although 10 patients had another type of brain lesion. We collected image-localized biopsies from 134 glioma surgeries, of which: 4 were grade 1, 16 were grade 2, 17 were grade 3 and 97 were grade 4. We collected image-localized biopsies from 38 surgeries for patients with cancers that had metastasized to the brain. See Fig 6 for further breakdowns such as tumor status and biopsy counts (analogous sex-specific figures can be found in S3 and S4 Figs).

**Fig 5:**
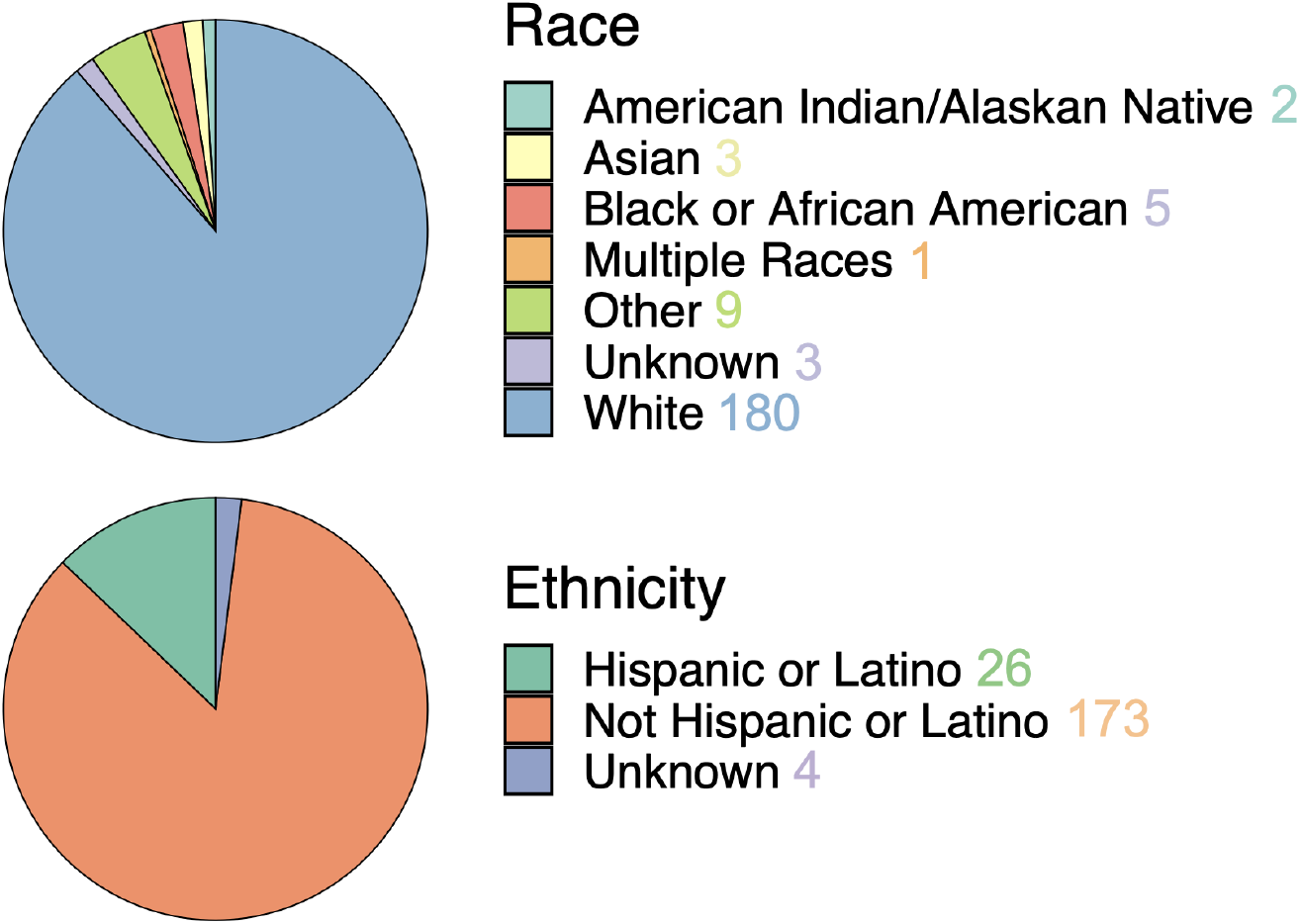
Self-reported race (top) and ethnicity (bottom) in the patient cohort, which consists predominantly of patients reporting as white and not Hispanic/Latino. See S2 Fig for sex-specific breakdowns.

**Fig 6:**
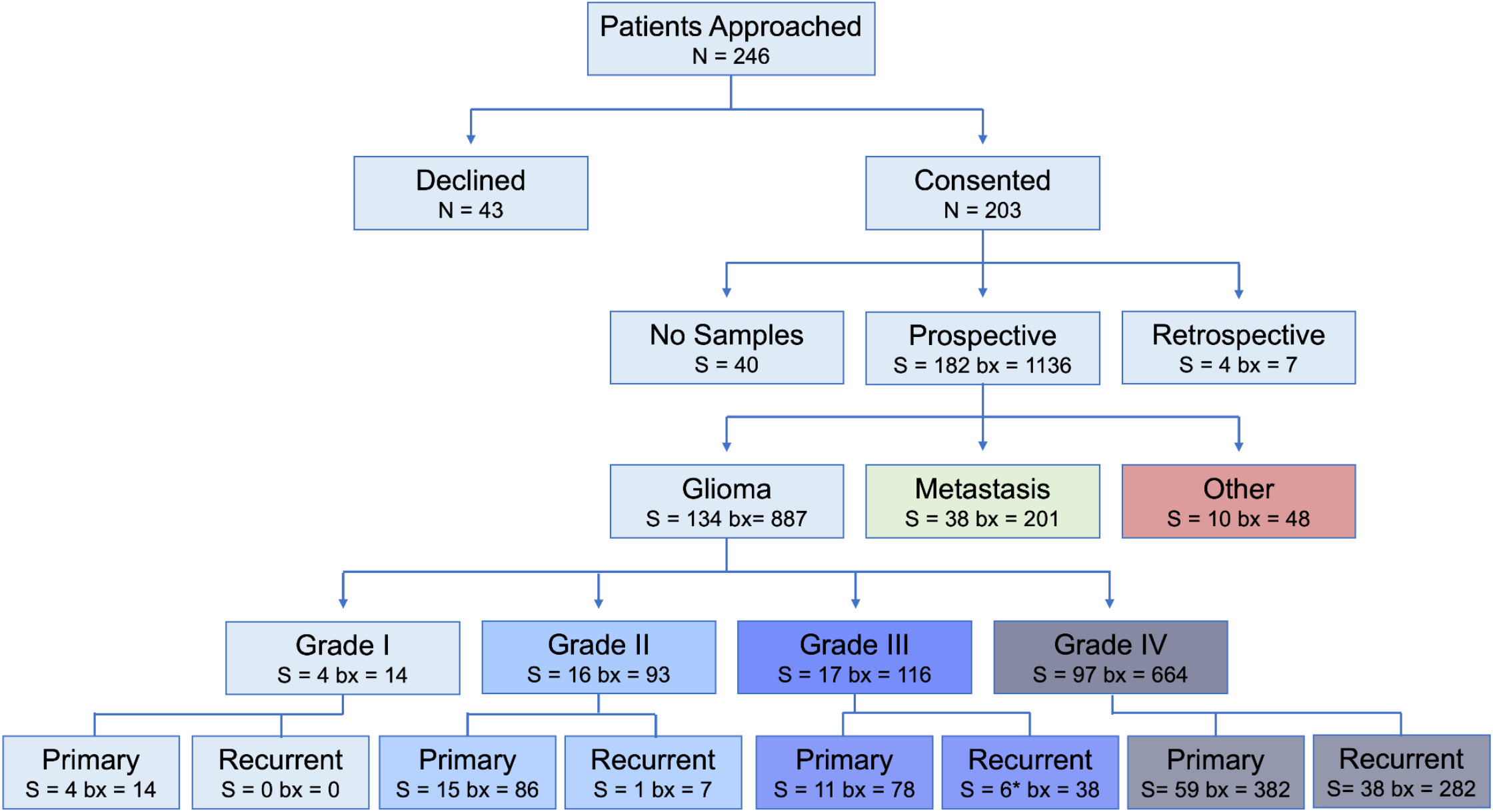
Flowchart for enrollment, starting from the number of patients (N) who were approached for the study, to surgeries (S) and total number of biopsies collected (bx), stratified by grade and tumor status. See S3 and S4 Figs for sex-specific breakdowns. *One case included in the recurrent grade III group underwent a grade transformation.

## Discussion

As with any clinical study, there are many potential limitations that must be considered. This study aims to retrieve multiple biopsies from a variety of locations spanning the diversity of MRI-defined tumor regions. For example in glioma, it is particularly important that we biopsy from the diffusely invaded normal appearing brain parenchyma, as these samples will be more similar to the unresected invasive margins that repopulate the disease. While we have a goal of 8 samples per surgery, in some cases this sampling scheme is not possible. In other cases, we may not be able to collect from all imaging regions. The operating neurosurgeon is responsible for evaluating the safety of biopsy collection and assessing any potential risk that may result from such collection. As a result, there is notable variability in the number and size of the collected specimens; however, this has helped define our study as a reasonable and pragmatic collection protocol.

Image-localization poses additional challenges. First, the neuronavigation system requires preoperative patient registration, resulting in a measurable registration error during surgery. Second, this process is reliant on preoperative imaging, which must be ordered by clinicians and, in some scenarios, is obtained up to three weeks prior to surgery. Further, such static images cannot account for intraoperative brain shift, a phenomenon where the brain fills the space of resected tissue. Unfortunately, this issue remains a universal challenge for image-localization during neurological surgery, and there is currently no standard procedure in place to accurately measure this change intraoperatively. These are all important considerations for image-based modeling. In addition, different MRI machines with different techs and field strengths can lead to varying imaging features, even within the same patient. Rather than alter clinical workflow, we must find ways (such as normalization techniques) to bring these images together into a comparable space for radiomics models.

Another limitation is in the uniformity of available imaging and the related difficulty of consenting patients promptly. Patients are identified through our institution’s patient scheduling systems (currently in EPIC, the electronic medical records system utilized at Mayo Clinic). Specifically, MRI and OR schedules are monitored for potential patients for the study. However, emergent patients who undergo imaging at short notice and immediately proceed with surgery may be missed or, if consented, may not have received the entire protocol of imaging before surgery. Since biopsy collections require research personnel, surgeries may also be missed if researchers’ schedules do not permit attendance. Further, surgeries are not attended if there is increased risk to the patient (e.g. patient safety concerns from the neurosurgeon) or the research team (i.e., active COVID-19 diagnosis).

Although this data collection comes with challenges, many of these are typical during the integration of research into a clinical workflow. Neuronavigation has proven to be a useful clinical tool for surgical planning and intraoperative guidance. By utilizing neuronavigation for research, we can attain much greater insight into the inter- and intra-patient spatial heterogeneity of brain tumors and its imaging presentation. These insights may provide therapeutic targets for clinical trials and we hope one day they will improve treatment options.

For all research approaches, we want our dataset to be both representative of each tumor and the patient cohort we ultimately aim to serve. Of course more data is always desirable, particularly for radiomics models, but it is unrealistic to expect the clinical team to collect all resected tissue as image-localized biopsies. With this protocol, we’ve mindfully struck a balance between representative data, minimal patient risk, and minimal impact on the clinical workflow. It is our hope that spatial localization of brain tumor biopsies becomes standard in the clinic to increase our understanding of these tumors, inform clinical care, and ultimately improve prognosis for patients.

## Data Availability

All data from tissue processed from this cohort will be made available through the MOSAIC consortium we have formed around this important novel intratumoral heterogeneity initiative: www.BrainTumorMOSAIC.org. Additional data requests may be initiated by contacting the Swanson lab through our website: www.MathematicalNeuroOncology.org.

## Authors’ Contributions

- Gustavo De Leon: manuscript conception
- Javier C.Urcuyo, Lee Curtin, Gustavo De Leon: manuscript preparation
- Javier C. Urcuyo, Lee Curtin, Gustavo De Leon, Jazlynn M. Langworthy: figure preparation, writing of manuscript and data management
- Barrett Anderies: initial development and deployment of biopsy protocol
- Javier C. Urcuyo, Lee Curtin, Jazlynn M. Langworthy, Gustavo De Leon, Barrett Anderies: Collection of tissue samples and biopsy coordinates
- Kyle W. Singleton, Andrea Hawkins-Daarud, Pamela R. Jackson, Sara Ranjbar: patient image processing
- Lisa Paulson: patient image analysis and segmentation of tumor abnormality on MRI
- Yvette Morris, Kamala Clark-Swanson: curation of clinical imaging and clinical data abstraction
- Chris Sereduk, Nhan L. Tran, Marcela Salomao, Kliment Donev: biopsy tissue processing and storage
- Andrea Hawkins-Daarud, Kamila M. Bond: post-processing biopsy data
- Maciej M. Mrugała, Alyx B. Porter, Leslie C. Baxter: patient identification and follow-up
- Miles Hudson, Jenna Meyer, Qazi Zeeshan, Mithun Sattur, Devi P. Patra, Breck A. Jones, Rudy J. Rahme, Matthew T. Neal, Naresh Patel, Pelagia Kouloumberis, Ali H. Turkmani, Mark Lyons, Chandan Krishna, Richard S. Zimmerman, Bernard R. Bendok: Patient identification and biopsy collection
- Barrett Anderies, Andrea Hawkins-Daarud, Pamela R. Jackson, Kristin R. Swanson: protocol development, establishment and management
- Leland S. Hu: manuscript review, development of imaging protocol, oversight of biopsies alignment, contributed to funding of biopsy team, oversight of image analysis, clinical PI of biopsy protocol
- Kristin R. Swanson: writing and review of manuscript, establishment of and management of overall biopsy protocol funding and team staffing, primary contributor of protocol funding and supervision of biopsy team, oversight of all protocol-related analyses, scientific PI of biopsy protocol

## Acknowledgements

One might say “it takes a village” to achieve a successful protocol of this scope. This manuscript focuses on the establishment of our protocol, current/recent workflows, processes, and staffing that we have implemented. As such, the authors are truly grateful to the many additional contributions by other prior and current members of the biopsy collection team, including but not limited to:

<u>Sample Collection:</u> Ariana Afshari, Spencer Bayless, Rhea Carlson, Ryan Hess, Julia Lorence, Reyna Patel, Lucas Paulson, Sebastian Velez, Ryan Eghlimi, Angad Beniwal, Gabe LaFond

<u>Clinical Research Coordinators:</u> Jessica Bauer, Regina Becker, Lysette Elsner, Crystal Harris, Morgan Hatlestead, Ashley Napier

<u>Surgical Technologists:</u> Darby Black, Heather Boles, Heather Hull, Angela Melloni, Jose Rocha, Briana Rodriguez

<u>Clinical Nursing Staff:</u> Nora J. Shaefer, Kaylee Curley, Hans Leitner DeCarlo, Deanna Dusek, Regina Formentin, Braden Hall, Nathan Nitzky, Melanie Parativo, Kenneth Rooth, Krystle Short, Matt Zumwalt Hulverson

<u>Stealth Techs:</u> Cara Burkholder, Albana Spahiu

<u>Interventional Radiology Technologists:</u> Jason Billings, Joel Dela Cruz, Emily Lange, Brandy Streeter, Patrick Vance

<u>Surgical Pathology Staff:</u> Bradley Atkins, Debra Keith, Phillip Hogan Jr, Rachel Matey, Lori Miller, Ashley Polzin, Jessica Williams

<u>Clinical data abstraction:</u> Sandra K. Johnston

<u>Clinical research database integration/administration:</u> Scott Whitmire

<u>Other project management and design:</u> Susan C. Massey

<u>Image analysis:</u> Cassandra R. Rickertsen and past and current members of the Image Analysis Team.

## Potential competing interests

KRS and LSH are co-founders of Precision Oncology Insights Inc; Imaging Biometrics (medical advisory board: LSH); the remaining authors have no relevant conflicts of interest to disclose. This does not alter our adherence to PLOS ONE policies on sharing data and materials.

## Funding statement

The authors gratefully acknowledge the funding that made this research possible from the NIH (U54CA143970, U54CA193489, R01CA16437, U01CA220378, U01CA250481), the NSF (1903135), the Zicarelli Foundation, the James S. McDonnell Foundation, the Ben and Catherine Ivy Foundation and Mayo Clinic.

## Notes

### Author Declarations

IRB of Mayo Clinic gave ethical approval for this work. IRB# 16-002424

### Summary of Updates

Updated to more recent sample counts.

