## Supplementary Material for "Image-localized biopsy mapping of brain tumor heterogeneity: A single-center study protocol"

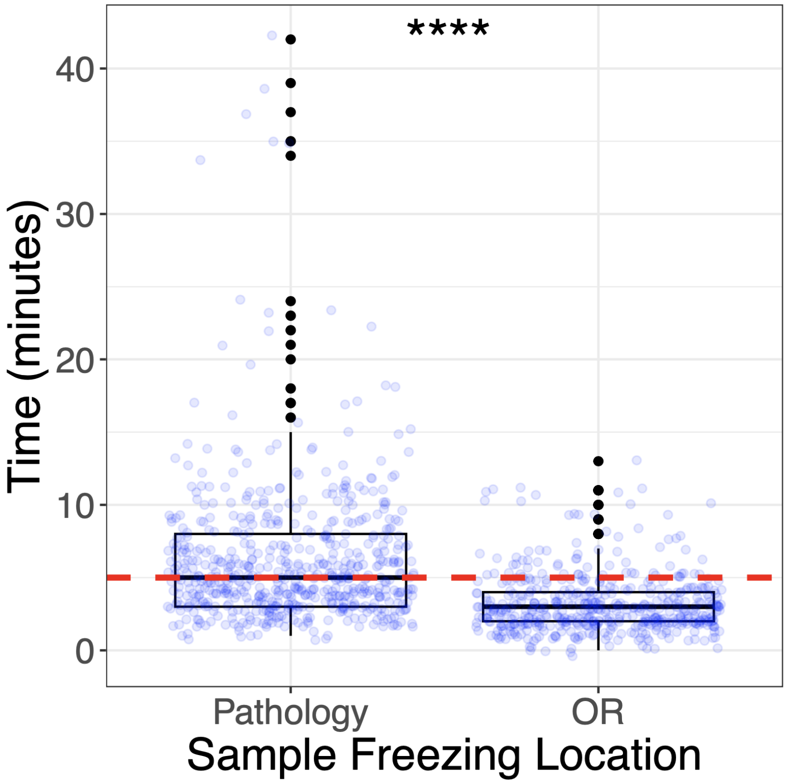


**S1 Fig. Time taken to freeze biopsy samples.** For 105/1126 of the flash-frozen samples, we have tracked the time from extraction to freezing. Switching from flash-freezing in pathology (N=597) to inside the operating room (OR, N=508) has allowed us to significantly reduce time between surgical extraction and freezing (linear mixed effects model with surgery ID as a nested random variable, p<0.0001). In doing so, we reach the goal of freezing biopsies in a 5-minute window (red dashed line) more consistently.

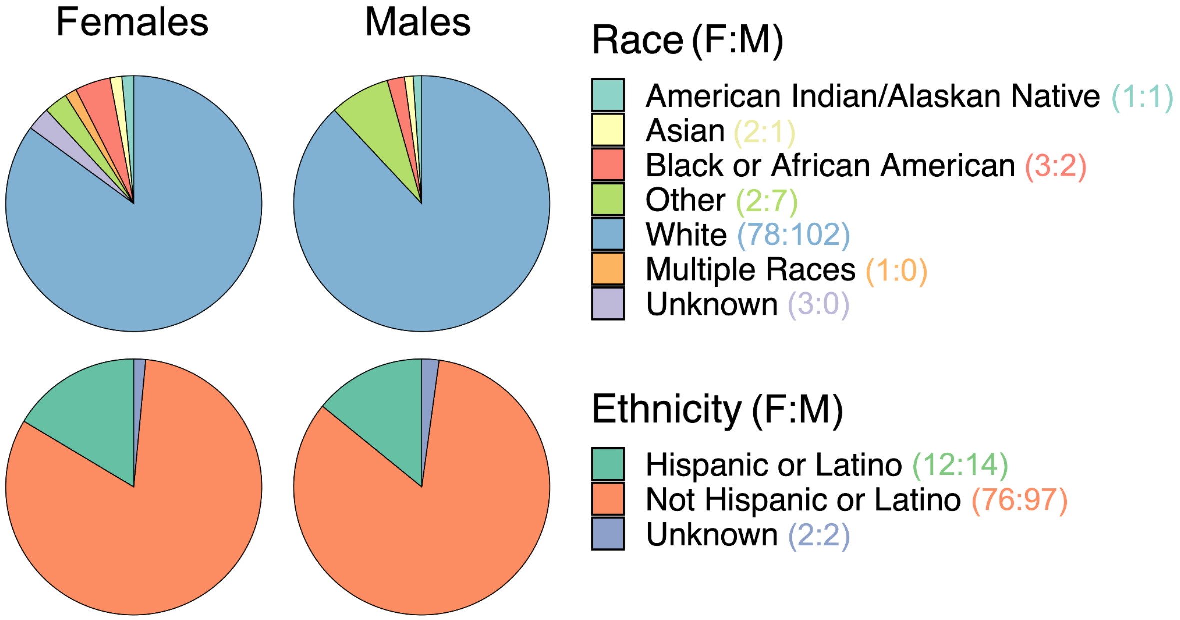


**S2 Fig. Self-reported race and ethnicity broken down by self-reported sex of patients.** Each sex consists predominantly of patients who identified as white, and patients who identified as not Hispanic/Latino.

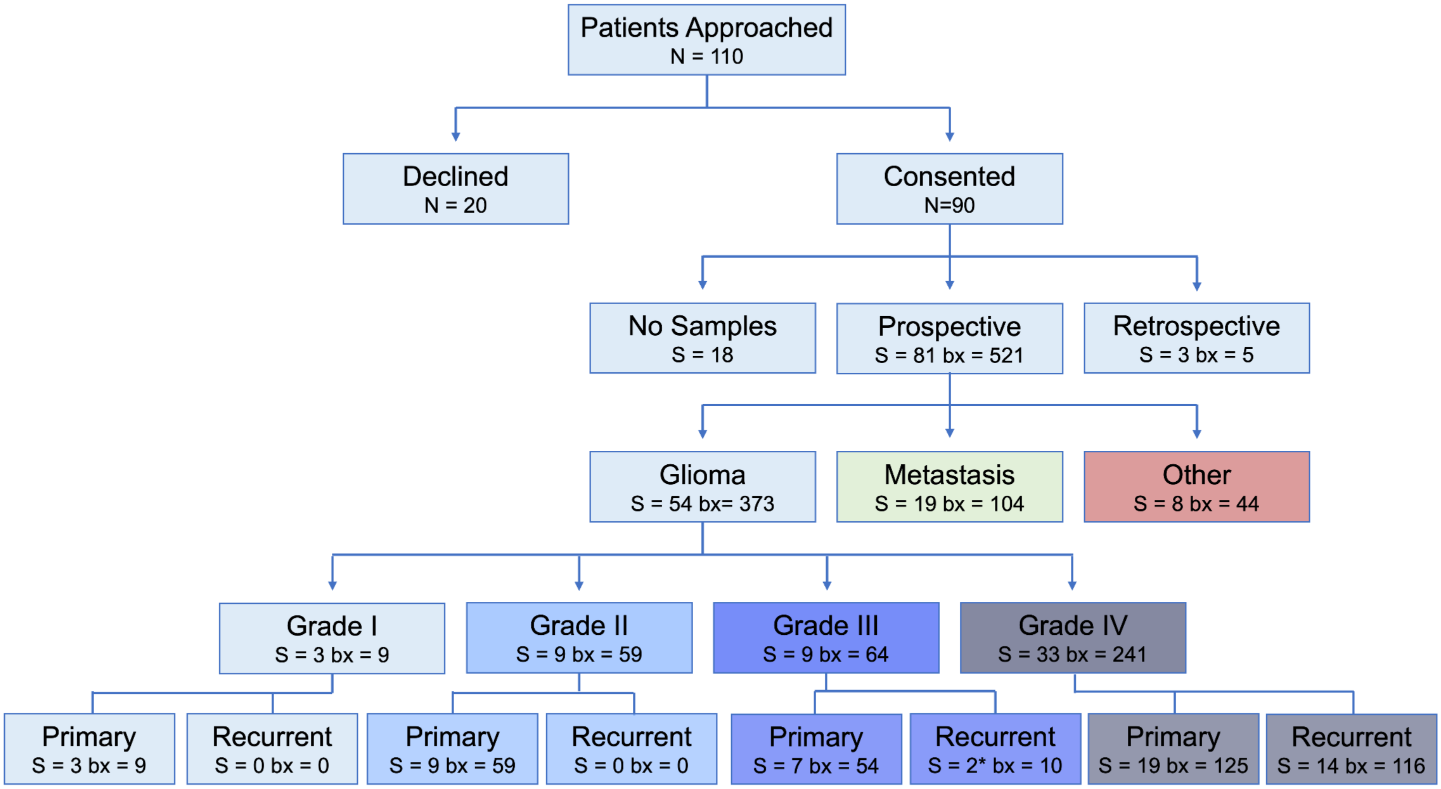


**S3 Fig. Counts breakdown for female patients**. The breakdown of patients approached and consented, as well as tumor type, grade and treatment status for patients who self-identified as female. *One case included in the recurrent grade III group underwent a grade transformation.

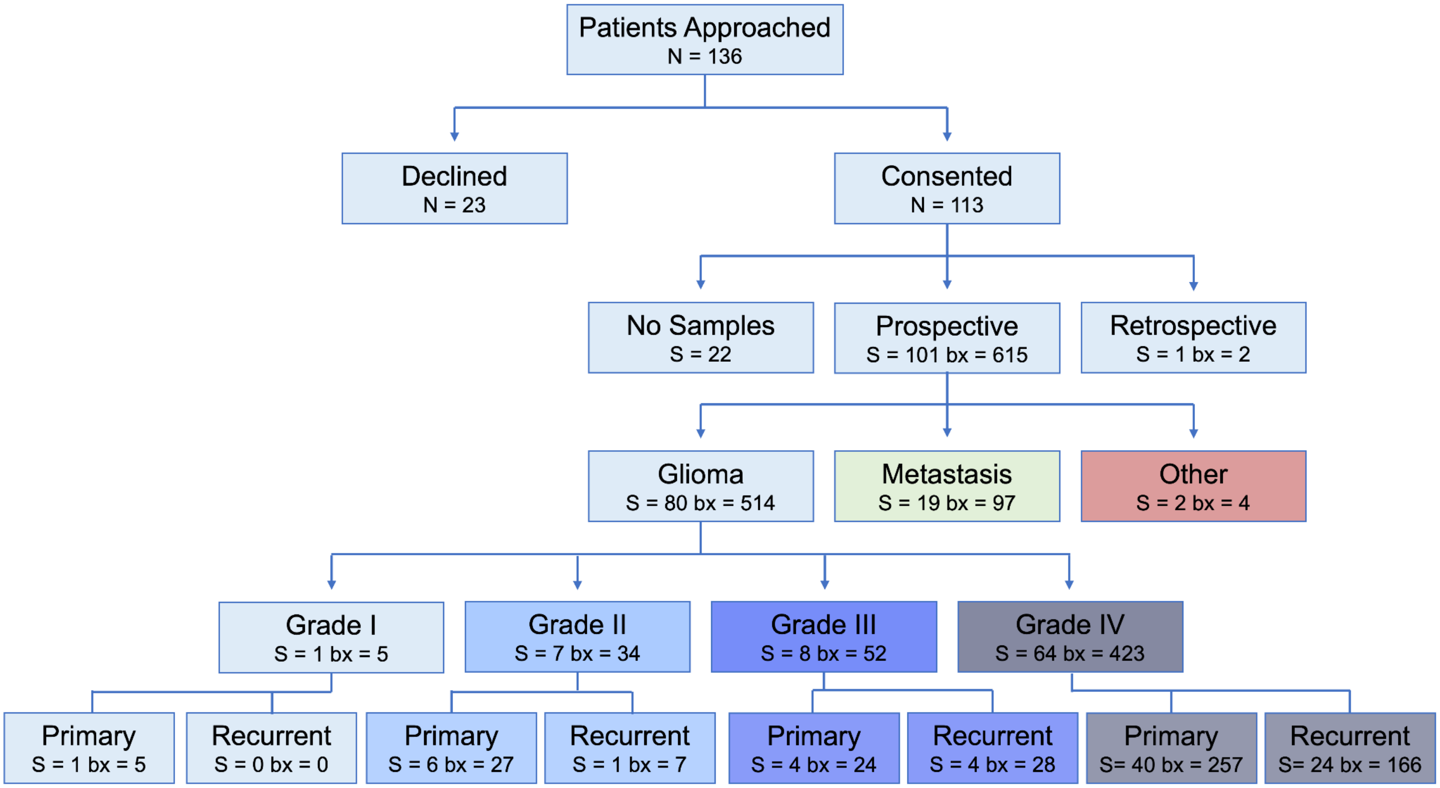


**S4 Fig. Counts breakdown for male patients**. The breakdown of patients approached and consented, as well as tumor type, grade and treatment status for patients who self-identified as male.
